# Acute psilocybin and ketanserin effects on cerebral blood flow: 5-HT2AR neuromodulation in healthy humans

**DOI:** 10.1101/2024.09.19.24313958

**Authors:** Kristian Larsen, Ulrich Lindberg, Brice Ozenne, Drummond E. McCulloch, Sophia Armand, Martin K. Madsen, Annette Johansen, Dea S. Stenbæk, Gitte M. Knudsen, Patrick M. Fisher

## Abstract

Psilocin, the active metabolite of psilocybin, is a psychedelic and agonist at the serotonin 2A receptor (5-HT2AR) that has shown positive therapeutic effects for brain disorders such as depression. To elucidate the brain effects of psilocybin, we directly compared the acute effects of 5-HT2AR agonist (psilocybin) and antagonist (ketanserin) on cerebral blood flow (CBF) using pseudo-continuous arterial spin labelling magnetic resonance imaging (MRI) in a single-blind, cross-over study in 28 healthy participants. We evaluated associations between plasma psilocin level (PPL) or subjective drug intensity (SDI) and CBF. We also evaluated drug effects on internal carotid artery (ICA) diameter using time-of-flight MRI angiography. PPL and SDI were significantly negatively associated with regional and global CBF (∼11.5% at peak drug effect, p<0.0001). CBF did not significantly change following ketanserin (2.3%, p=0.35). Psilocybin induced a significantly greater decrease in CBF compared to ketanserin in the parietal cortex (p_FWER_<0.0001). ICA diameter was significantly decreased following psilocybin (10.5%, p<0.0001) but not ketanserin (−0.02%, p=0.99). Our data support an asymmetric 5-HT2AR modulatory effect on CBF and provide the first in vivo human evidence that psilocybin constricts the ICA, which has important implications for understanding the neurophysiological mechanisms underlying its acute effects.

## Introduction

Psilocybin has emerged as a promising therapeutic showing rapid and lasting reductions in clinical symptoms across several brain disorders, including major depression,^1–3^ cancer related anxiety and depressive symptoms,^4,5^ and cigarette and alcohol addiction. ^6^ A psychedelic dose of psilocybin induces subjective effects lasting 6-8 hours characterised by visual alterations, profound mood changes and shifts in the perception of space, time and selfhood.^7,8^ Psilocybin has also been shown to produce lasting changes in self-reported measures including increased personality trait openness, decreased neuroticism, increased mindfulness and life-satisfaction including in healthy individuals.^9–16^ After intake, psilocybin is dephosphorylated to its active metabolite, psilocin (4-OH-N,N-dimethyltryptamine),^17^ which is a partial agonist at several serotonergic receptors, including the serotonin 2A receptor (5-HT2AR). ^18^ Accumulating evidence shows 5-HT2AR stimulation underlies the acute psychedelic effects ^19–24^, e.g., pre-treatment with ketanserin blocks the acute psychedelic effect of psilocybin,^20^ and other classical psychedelics,^21^ and ketanserin administration reverses acute psychedelic effects following lysergic acid diethylamide (LSD).^25^ Neuronal 5-HT2ARs are predominantly expressed on the apical dendrites of excitatory pyramidal neurons in the cortical layers III and V, GABAergic neurons, and monoaminergic terminals.^26–28^ Notably, 5-HT2ARs are also expressed on smooth muscle cells in blood vessels, where they can induce vasoconstriction and consequently increased blood pressure.^29–31^ In vitro studies confirm that 5-HT2AR-induced arterial contractions are abolished by antagonists like ketanserin, which acts as a vasodilator in the presence of serotonin.^32–36^ Understanding the *in vivo* neurovascular effects of 5-HT2AR modulation could enhance our knowledge of how psychedelics affect brain function.

Recent blood oxygen level-dependent functional magnetic resonance imaging (BOLD fMRI) and electroencephalography (EEG) studies of human brain function show substantial acute effects of psychedelics on functional brain connectivity. Most consistently, resting-state BOLD fMRI scans during the acute experience increases global connectivity, particularly between transmodal resting-state networks. ^37,38^ Estimating CBF with arterial spin labelling (ASL) can provide complementary information related to neural signalling that can be estimated in absolute units (ml/100g/min) and may inform neural correlates of psychedelic drug effects.^39^ Acute psilocybin effects on CBF have been previously reported in three studies using ASL fMRI.^40–42^ Carhart-Harris and colleagues^40^ reported decreased CBF up to 12 minutes following intravenous administration of 1-2 mg of psilocybin in 15 healthy participants. Lewis and colleagues^41^ reported decreased CBF approximately 60 minutes following oral psilocybin administration of 0.16 or 0.215 mg/kg in 58 healthy participants, an effect echoed in a follow-up analysis including 12 additional participants^42^. Lewis and colleagues also considered an alternative CBF quantification strategy using regional relative hyperperfusion, which they argued aligns with results following psychedelic administration using single photon emission computed tomography (SPECT) and positron emission tomography (PET) studies of CBF and glucose metabolism, respectively.^43,44^ Contrasting acute psilocybin effects with a 5-HT2AR antagonist would contribute to our understanding of how 5-HT2AR modulation shapes neurovascular function.

In this randomised, single-blind, cross-over study in healthy participants, we contrasted the acute effects of a single oral dose of psilocybin or ketanserin on CBF with pseudo-continuous ASL (pcASL). We mapped changes in CBF at several time points after psilocybin administration. To more comprehensively characterise cerebrovascular effects, we examined acute drug effects on internal carotid artery (ICA) diameter. We evaluated the association between CBF and plasma psilocin levels (PPL) and subjective drug intensity (SDI), which are related to brain 5-HT2AR receptor occupancy and functional connectivity.^45,46^

## Methods and materials

### Participants

Twenty-eight healthy volunteers participated in this study (10 females, age (mean ± SD): 33 ± 8 years). Participants were recruited from a database of individuals interested in participating in a brain imaging study involving the administration of psychedelics. BOLD fMRI data unrelated to the current study and from a subset of participants described herein has been reported in previous studies.^7,16,46–48^ Here, we present the pcASL data for the first time. Recruitment began in September 2018 and ended in March 2021. Forty-four individuals were assessed for eligibility, of which 16 were excluded (see Figure 1). Exclusion criteria were 1) present or previous personal or family (first degree relatives) history of psychiatric disease (DSM axis 1 or WHO ICD-10 diagnostic classification) including substance misuse disorder; 2) present or previous neurological condition/disease, significant medical disorders or intake of drugs suspected to influence test results; 3) non-fluent in Danish; 4) vision or hearing impairment; 5) learning disability; 6) pregnancy; 7) breastfeeding; 8) MRI contraindications; 9) alcohol or drug abuse; 10) allergy to test drugs; 11) significant exposure to radiation within the past year (e.g., medical imaging investigations); 12) intake of QT-prolonging medication or electrocardiogram (ECG) results indicative of heart disease; 13) history of significant adverse response to a hallucinogenic drug; 14) use of hallucinogenic drugs less than 6 months prior to inclusion; 15) blood donation less than 3 months before project participation; 16) body weight less than 50 kg; 17) low plasma ferritin levels (< 12 μg/L). Standard laboratory blood sample tests were conducted and an electrocardiogram was taken to ensure that individuals were in good overall physiological health (e.g., absent evidence for a prolonged QT interval). Prior to inclusion, participants received written and verbal descriptions of the study protocol, including the expected effects of psilocybin and potential side effects of psilocybin and ketanserin; all participants provided written consent prior to study participation. The data presented here is part of a broader study approved by the ethics committee for the Capital Region of Copenhagen (journal identifier: H-16028698, amendments: 56023, 56967, 57974, 59673, 60437, 62255) and Danish Medicines Agency (EudraCT identifier: 2016-004000-61, amendments: 2017014166, 2017082837, 2018023295).

**Figure 1:**
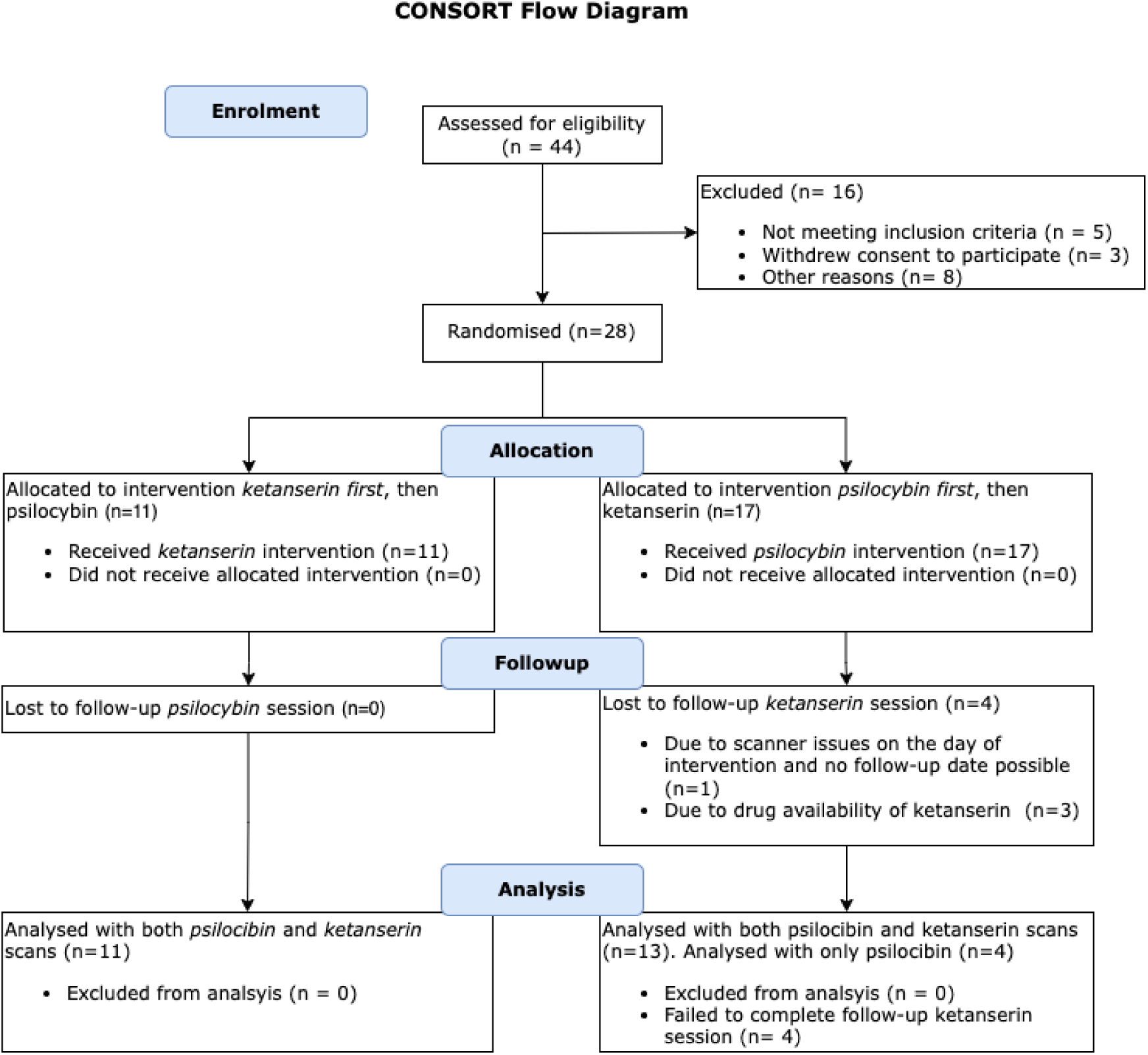
CONSORT Flow Diagram. This diagram shows the enrollment and allocation of participants in a study evaluating the effects of psilocybin and ketanserin on cerebral blood flow. 44 participants were assessed for eligibility, with 28 being randomised into the study. 17 participants received psilocybin first and 11 received ketanserin first, with a minimum 3 week washout period in between. All 28 participants completed the psilocybin intervention, whereas 4 participants failed to complete the ketanserin session.

### Study design

This study used a single-blind, within-subject, cross-over design with two drug intervention sessions: (i) psilocybin and (ii) ketanserin. Seventeen participants were assigned to receive psilocybin first, 11 assigned to ketanserin first. The allocation was generated by a staff member who was not otherwise involved in the project. Drug sessions were separated by at least three weeks (mean: 31 days, median: days, range: [21-105]; time between sessions was more than 42 days for only one participant due to delay following the Covid-19 lockdown). Additional details about the overall study design have been described previously.^46^

### Study procedures

Participants were asked to refrain from alcohol the night before intervention days, to eat a light breakfast and to refrain from caffeine intake on the day of intervention. Before drug administration and pcASL data acquisition, a urine sample was obtained and screened for amphetamines, opioids, benzodiazepines, barbiturates, tetrahydrocannabinol, cocaine, ketamine, phencyclidine, and gamma-hydroxybutyrate (Rapid Response, BTNX Inc). Participants met with two psychologists before the intervention day for preparation (approx. one week) and again the day after the psilocybin intervention for integration of the experience. On intervention days, between scans, participants retired to a comfortable and pleasant room adjacent to the MR scanner, where music was played. At least one psychologist was present with the participant to provide interpersonal support throughout the psilocybin intervention, including during MR scanning.

Participants completed a pre-drug MRI scan including structural, pcASL, and angiography scans (detailed below). The pre-drug scan was followed by a brief conversation with the psychologists, after which drug was administered. We administered psilocybin according to body weight in units of 3 mg capsules (0.26 ± 0.04 mg/kg, absolute dose 19.8 ± 3.7 mg, mean ± SD) and ketanserin as a single 20 mg tablet. These doses were selected to achieve a comparable peak 5-HT2AR occupancy 1-3 hours after administration of approximately 70% (see Supplementary Figure S1).^45,49,50^ Drugs were given with a glass of water. On psilocybin days, pcASL scans were acquired at approximately 40, 80, 130, and 300 minutes post-drug to capture both peak drug effects as well as the subsequent decline. Blood samples for PPL and SDI assessments were taken after each scan. Participants were sent home once SDI levels dropped below 1 after the final scan.On ketanserin days, pcASL scans were acquired before and at approximately 80 minutes post-drug. While not in the scanner, participants were listening to music in the adjacent room. On ketanserin days, participants with an SDI < 1 after 130 minutes completed self-report questionnaires and were sent home following a short debriefing.

### Plasma psilocin level

PPL was measured in venous blood samples drawn from a catheter in the antecubital vein after each pcASL scan. The blood samples were centrifuged and the plasma fraction frozen and briefly stored at −20°C before being transferred to −80°C until analysis. Plasma psilocin levels were measured using ultra performance liquid chromatography and tandem mass spectrometry, as previously described,^45^ reflecting free, unconjugated psilocin and not glucuronidated psilocin.^51^ Plasma psilocin is reported in units μg psilocin per L plasma (μg/L).

### Subjective drug intensity

Following each pcASL scan, an SDI rating was acquired by asking the participant: “How intense is your experience right now on a scale from 0 to 10?” (Likert scale, 0 = not intense at all, 10 = very intense).

### Magnetic resonance imaging

Participants were scanned on one of two 3 Tesla Siemens Magnetom Prisma MRI scanners (Siemens AG, Erlangen, Germany) at Rigshospitalet, Copenhagen (MR1 and MR2). Each participant completed all their scans on only one MRI scanner. Fifteen participants completed scan sessions on MR1 and thirteen participants on MR2. All images were visually inspected for quality using the FSL image viewer, FSLeyes.^52^

### Magnetic resonance imaging parameters

#### MR1

Related data from the participants scanned on this scanner has been reported previously.^46,47^ Imaging data were acquired using a 64-channel head/neck coil. During the pre-drug scan we acquired a high-resolution, T1-weighted 3D MPRAGE structural image (inversion time = 900ms, echo time = 2.58ms, repetition time = 1900ms, flip angle = 9°, in-plane matrix = 256×256, in-plane resolution = 0.9mm×0.9mm, number of slices: 224, slice thickness = 0.9mm, no gap). CBF was estimated using a 2D echo-planar imaging (EPI) gradient spin-echo sequence (scan time = 5.33 min, echo time = 12ms, repetition time = 4000ms, flip angle = 90°, label duration: 1508ms, single post-labeling delay = 1500ms, in-plane matrix = 64×64, in-plane resolution = 3mm×3mm, number of slices = 20, slice thickness = 5mm no gap, number of control/label pairs = 40, slice readout duration = 35ms). To calibrate the pcASL signal, an M0 scan was acquired using the same imaging parameters as the pcASL except for a repetition time of 10000ms. We acquired a gradient echo sequence (GRE) to correct for geometric distortions in the pcASL images (echo time 1 = 4.92ms, echo time 2 = 7.38ms, repetition time = 400ms).

#### MR2

Imaging data were acquired using a 32-channel head coil. During the pre-drug scan, we acquired a high-resolution, T1-weighted 3D MPRAGE structural image (inversion time = 920ms, echo time = 2.41ms, repetition time = 1810ms, flip angle = 9°, in-plane matrix = 288×288, in-plane resolution = 0.8×0.8mm^2^, slice thickness = 0.8mm, number of slices = 224, no gap). CBF was estimated using a five post-label delay 3D turbo gradient spin echo sequence (scan time = 7.11min, 12 label/control pairs, native voxel size: 2.5×2.5×3mm^3^, image matrix = 96×96×40, label duration = 1508ms, post-label delays = [500, 500, 1000, 1000, 1500, 1500, 2000, 2000, 2000, 2500, 2500, 2500]ms, echo time = 3.78ms, repetition time = 4100ms, flip angle: 120°), two background suppression pulses optimised for each PLD.. The acquisition included an M0 scan with which to calibrate the pcASL signal without any background suppression. Spin-echo echo-planar acquisitions with opposite phase-encoded blips were acquired to correct for geometric distortions using *topup* implemented in FSL (FMRIB software library, www.fmrib.ox.ac.uk).^53,54^ Notably, for the 13 participants scanned on MR2, we also acquired a time-of-flight (TOF) MR angiography sequence using a 3D flow-compensated gradient echo technique with multiple slabs (echo time = 3.42ms, repetition time = 19ms, flip angle = 18°, slice thickness = 1.3mm, spacing between slices = 1.3mm, base resolution = 256×256, phase resolution = 0.8, partial Fourier factor = 0.75).

### Participant exclusion and missing data

During psilocybin intervention days, four participants did not complete all planned pcASL scans. On MR1, one participant missed the post-drug scan at +300-min due to technical issues, and another did not complete it due to nausea. On MR2, one participant did not complete any post-drug scans due to discomfort, while another missed the +130-min and +300-min scans due to nausea. For the ketanserin session, four participants were excluded: one due to the Covid-19 lockdown, and three due to drug availability issues. We analysed only the pre-drug and +80-min scans, which corresponds to periods of comparable 5-HT2AR occupancy between psilocybin and ketanserin (see Supplementary Figure S1).^45,49^

### pcASL data quality control

Several scans were excluded due to quality issues. Two post-psilocybin scans from one MR1 participant were excluded due to excessive head motion. Additionally, five post-psilocybin scans (four from MR1 and one from MR2) were excluded for the same reason. On ketanserin days, four scans from two MR2 participants were excluded due to image artefacts, and three additional scans were excluded for falling outside the peak occupancy time window.

### CBF quantification

#### MR1 preprocessing

The pcASL data underwent intramodal registration dividing label and control images (40 pairs) before realigning them. The pcASL data were then unwarped to correct for B0 field distortions. Realignment and distortion correction was then performed using SPM12 (https://www.fil.ion.ucl.ac.uk/spm/software/spm12/). The high-resolution T1-weighted structural image was co-registered with the realigned and motion corrected pcASL data. We corrected for the interleaved slice acquisitions in the 2D EPI protocol using a delay in slice time acquisition of 35ms.

#### MR2 preprocessing

For each of the PLDs, a label/control difference image was realigned and the high-resolution T1-weighted structural image was co-registered with the M0 image. Co-registration with the pcASL images like we did for the MR1 data was not possible without introducing transformation artefacts.

All pcASL images from both scanners were quantified to absolute CBF values using BASIL in FSL (FMRIB software library, www.fmrib.ox.ac.uk),^55^ which performs subtraction of label/control images and calibrates the signal to absolute CBF (ml/100g/min) incorporating the M0 scan and modelling the tracer kinetics using a single, well-mixed tissue compartment model.^56^ We used standard settings for the quantitative modelling described previously.^56^ The structural image was segmented in SPM12 into grey matter, white matter, and cerebrospinal fluid probability maps. The calibrated pcASL image was normalised into (MNI) standard space based on estimated warps from the segmentation step. Normalised pcASL images were spatially smoothed with a 3D Gaussian kernel of 8 mm Full-Width at Half-Maximum (FWHM).

### ICA diameter quantification

We developed and applied an in-house, semi-automatic tool (GitHub link: https://github.com/kristian1801/ica-segmentation-tool) for ICA segmentation and diameter quantification. TOF images underwent preprocessing to enhance vascular structures, including background subtraction, anisotropic diffusion filtering, and top-hat filtering. User-guided segmentation was performed via a graphical interface, where the user manually selected the ICA. A 2D region-growing algorithm with dynamic intensity thresholding was then applied to segment the chosen vessel across all axial slices. The software reconstructed a 3D model of the segmented ICA and calculated its diameter by taking the median of measurements from each image slice.

## Data analysis

### Regions of interest

Regions of interest (ROIs) for CBF-associated analyses were defined using the Anatomical Automatic Labelling (AAL3) atlas.^57^ The AAL3 delineates 170 ROIs in MNI space but here we grouped adjacent ROIs to form 13 larger, bilateral ROIs for the analysis, based on previous studies of acute psilocybin effects on CBF ^40,41^: prefrontal cortex, temporal cortex, parietal cortex, occipital cortex, anterior cingulate cortex (ACC), posterior cingulate cortex (PCC), thalamus, amygdala, putamen, caudate nucleus, hippocampus, insula, orbitofrontal cortex (OFC). See Supplemental Material for a description of AAL3 regions combined to produce each ROI.

We computed and evaluated grey-matter weighted regional CBF as defined by subject-specific grey-matter segmentation maps. We computed global grey-matter weighted CBF across all voxels within cortical and subcortical regions in the AAL3 atlas. The cerebellar ROI was excluded from analyses due to inconsistent field of view coverage across participants.

### Associations of CBF and ICA diameter with PPL and SDI

A total of 124 pcASL scans were included from psilocybin intervention days (70 from MR1, 54 from MR2), and 49 pcASL scans were included from ketanserin days (30 from MR1, 19 from MR2). We used linear mixed-effects models (LME) to estimate associations between exposure variables (PPL or SDI) and brain measurements (global or regional CBF, or ICA diameter). We included participant ID as a random effect to account for correlations between repeated observations from the same participant; age, sex, and scanner were included as fixed effects. We fitted separate LMEs for each combination of outcome (global CBF, 13 regional CBF measurements, and ICA diameter) and exposure (PPL or SDI). The regression coefficient β quantifies the association, representing the average change in the brain measurement (Y) for a one-unit increase in the exposure variable (X), while keeping other characteristics constant. We assessed statistical significance using Wald tests. For the 13 regional CBF tests, we adjusted p-values using Dunnett’s method to control the family-wise error rate (p_FWER_). ^58^ We did not adjust p-values for global CBF and ICA diameter. We used a statistical significance threshold of p or p_FWER_ < 0.05. We also report the estimated maximum percent change as 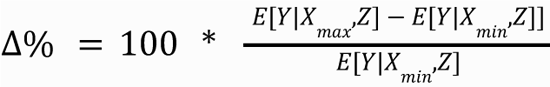, where E[Y|X, Z] denotes the expected value of the brain measurement Y, given the exposure X and other covariates Z, X_max is the maximum exposure value (19.3 μg/L for PPL, 10 for SDI) and X_min is the minimum exposure value (0 for both PPL and SDI). As an exploratory analysis, we estimated receptor occupancy using PPL values and the Emax model from our previous work ^45^ (Supplementary Figure S2): Occupancy = (Occ_max_* PPL) / (EC_50_ + PPL), where EC_50_ is the concentration at half-maximal effect. Occ_max_ was set as 76.6% and EC_50_ as 1.96 µg/L.

### Ketanserin effects and drug-x-time effects on CBF and ICA diameter

For ketanserin effects, each model included time (pre-drug vs. +80-min) as the independent variable, with sex, age, and scanner as covariates. We reported percent change in CBF and ICA following ketanserin administration using the same formula as described following formula: 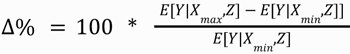, denotes the expected value of the brain measurement Y, given the exposure X and other covariates Z. X_max represents the peak time point (+80-min post-drug administration), and X_min represents the pre-drug timepoint. Drug-x-time interaction effects models were structured similarly to the ketanserin effects models, with the addition of a drug-x-time interaction term, allowing for comparison of the effect of psilocybin versus ketanserin from pre-drug to +80-min. For a comparison of the occupancy estimates of psilocybin and ketanserin, please refer to supplementary Figure S1. We adjusted p-values using Dunnett’s method to control the family-wise error rate (p_FWER_). We did not adjust P-values for global CBF and ICA diameter. We evaluated all associations using a statistical significance threshold of p or p_FWER_ < 0.05.

### Visualisations, code and data availability

The MATLAB and R code used to generate the results presented in this study can be made available upon request. We used BrainNet Viewer^59^ (https://www.nitrc.org/projects/bnv/) to generate the brain colour maps presented in Figure 2a. The datasets generated and/or analysed during the current study can be made available upon completion of a formal data-sharing agreement.

**Figure 2.**
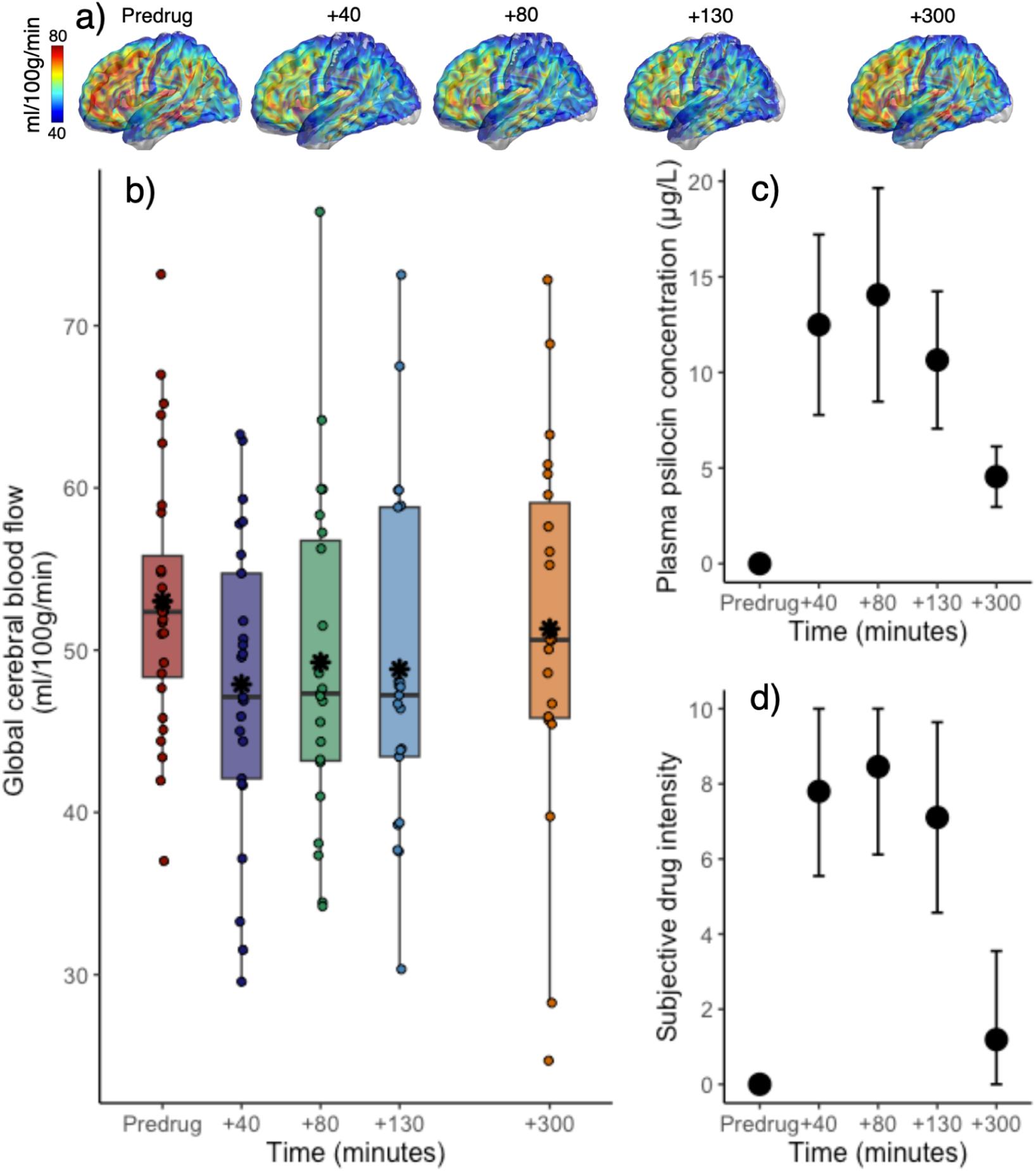
Time course of cerebral blood flow (CBF), plasma psilocin level (PPL) and subjective drug intensity (SDI) before and after psilocybin intake. **a)** Brain representations are colour maps created from the absolute voxel-wise CBF values. b) A boxplot of global, grey-matter weighted CBF at pre-drug, +40-, +80-, +130- and +300-min following psilocybin administration. Asterisks mark the mean value. b) and c) temporal trajectories of PPL and SDI (group mean ± 1 SD) associated with imaging sessions.

## Results

On psilocybin days, we observed ranges for PPL [0 - 27.6 μg/L], SDI [0 - 10], and CBF [22.9 - 77.0 ml/100g/min] throughout the time course of the intervention. The time course of PPL and SDI over the experiment was similar, confirming both drug uptake and associated subjective effects (Figure 2b-c). All participants reported psychoactive effects following psilocybin administration, as evidenced by high ratings on the SDI scale. In contrast, ketanserin produced negligible subjective effects, and most participants rated their SDI at or near zero at the +80-min post-drug scan session (SDI score: mean = 0.23, median = 0, range = 0-3).

### Global CBF decreases following psilocybin intake

We observed decreased global CBF following psilocybin administration (Figure 2a). There was a significant negative association between global CBF and both PPL (ß=-0.38, 95% CI = [−0.56, −0.20] ml/100g/min per μg/L of psilocin, p < 1e-04) and SDI (ß=-0.75, 95% CI = [−0.1, −0.41] ml/100g/min per unit SDI, p < 1e-04), see also Table 1 and Figure 3. This corresponds to a ∼11.6% decrease in global CBF at peak PPL or SDI.

**Figure 3.**
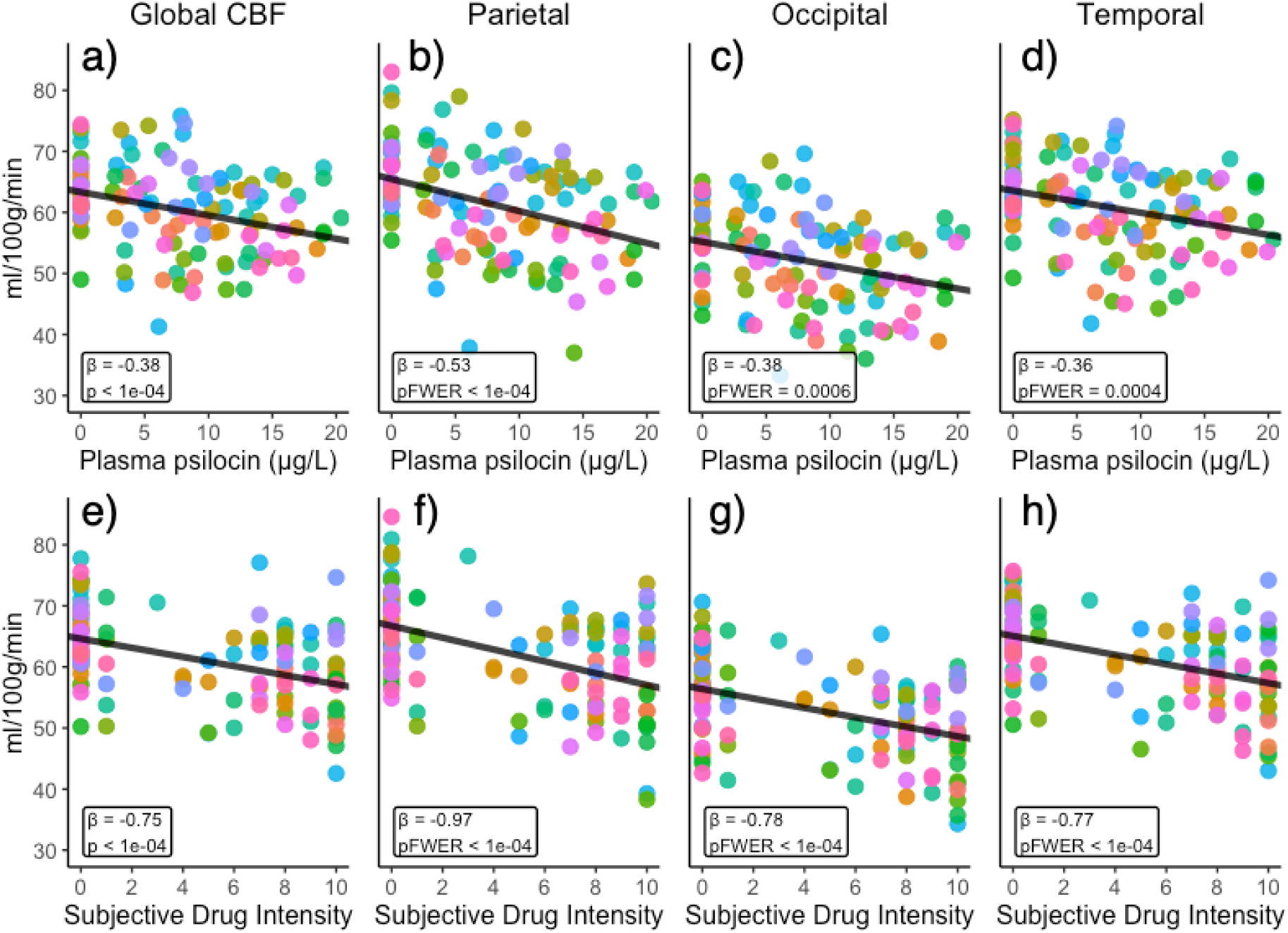
Associations between regional cerebral blood flow (CBF) and plasma psilocin level (PPL) and subjective drug intensity (SDI). **a), b), c)** and d) display the global, parietal, occipital and temporal region associations with PPL, respectively. Panels e), f), g) and h) display associations with SDI. Associations with CBF are plotted adjusting for age, sex and scanner. Each colour denotes a unique individual.

**Table 1.** Associations with plasma psilocin level and subjective drug intensity.

| MEASURE | Plasma psilocin level |  |  |  | Subjective drug intensity |  |  |  |
| --- | --- | --- | --- | --- | --- | --- | --- | --- |
| | $\beta$ | SE | $P_{FWER}$ | $\Delta\%$ | $\beta$ | SE | $P_{FWER}$ | $\Delta\%$ |
| **Global CBF** | **-0.38** | **0.09** | **<1e-04\*\*\*** | **-11.62** | **-0.75** | **0.12** | **<1e-04\*\*\*** | **-11.56** |
| Parietal | -0.53 | 0.10 | <1e-04*** | -15.47 | -0.97 | 0.17 | <1e-04*** | -14.51 |
| Occipital | -0.38 | 0.10 | 0.0006*** | -13.40 | -0.78 | 0.15 | <1e-04*** | -13.77 |
| Prefrontal | -0.30 | 0.10 | 0.017* | -8.22 | -0.58 | 0.15 | 0.0008*** | -8.25 |
| Temporal | -0.36 | 0.09 | 0.0004*** | -11.00 | -0.77 | 0.16 | <1e-04*** | -11.86 |
| OFC | -0.04 | 0.13 | 1.00 | -1.20 | -0.23 | 0.24 | 0.88 | -3.67 |
| ACC | -0.33 | 0.11 | 0.017* | -8.58 | -0.70 | 0.19 | 0.001** | -9.18 |
| PCC | -0.59 | 0.16 | 0.003** | -13.88 | -1.22 | 0.27 | <1e-04*** | -14.52 |
| Insula | -0.22 | 0.13 | 0.43 | -5.96 | -0.57 | 0.24 | 0.10 | -7.95 |
| Putamen | -0.15 | 0.09 | 0.43 | -5.55 | -0.54 | 0.16 | 0.008** | -9.83 |
| Caudate | -0.11 | 0.10 | 0.79 | -4.79 | -0.35 | 0.16 | 0.14 | -7.45 |
| Thalamus | -0.03 | 0.14 | 1.00 | -1.13 | -0.40 | 0.25 | 0.44 | -7.84 |
| Hippocampus | 0.03 | 0.11 | 1.00 | 1.03 | -0.30 | 0.21 | 0.55 | -5.17 |
| Amygdala | -0.23 | 0.12 | 0.29 | -9.29 | -0.51 | 0.23 | 0.15 | -10.57 |
| **ICA diameter** | **-0.02** | **0.003** | **<1e-04\*\*\*** | **-10.51** | **-0.03** | **0.01** | **<1e-04\*\*\*** | **-6.61** |

Expected change in outcome measure (cerebral blood flow or internal carotid diameter) for a unit change in plasma psilocin level (PPL) or subjective drug intensity (SDI) as estimated by the linear mixed model. $\beta$ represents the estimated change. $\Delta\%$ denotes the estimated percent change for PPL = 19.3 $\mu\text{g/L}$ (max value based on previous study [49]) versus PPL = 0, and SDI = 10 (max value) versus SDI = 0. $P_{FWER}$ indicates family-wise error rate corrected p-values, except for Global CBF and ICA diameter, which are uncorrected. CBF measures are reported for global and regional areas. Brain regions include OFC (Orbitofrontal cortex), ACC (Anterior cingulate cortex), and PCC (Posterior cingulate cortex). SE: standard error. Significance levels: \* $p < 0.05$ , \*\* $p < 0.01$ , \*\*\* $p < 0.001$ .

### Associations between CBF and PPL or SDI

All regions, except the orbitofrontal cortex (OFC) and hippocampus, displayed a numerically negative association between PPL and CBF (Table 1). The effects were statistically significant in the parietal, occipital, All regions displayed a numerically negative association between SDI and CBF (Table 1). Effects were estimated receptor occupancy and CBF (Supplementary Figure S2 and Table S1).

Intake of ketanserin was not associated with a statistically significant change in global CBF (p = 0.35, Table 2), Intake of ketanserin was not associated with a statistically significant change in ICA diameter (p = 0.99), see Table 2.

**Table 2.** Ketanserin and drug-x-time effects.

| MEASURE | Ketanserin effect |  |  |  | Drug-x-time |  |  |  |
| --- | --- | --- | --- | --- | --- | --- | --- | --- |
| | $\beta$ | SE | $P_{FWER}$ | $\Delta\%$ | $\beta$ | SE | $P_{FWER}$ | $\Delta\%$ |
| **Global CBF** | **-2.05** | **1.05** | **0.35** | **2.30** | **-3.54** | **2.38** | **0.14** | **-5.09** |
| Parietal | -0.84 | 0.92 | 0.95 | 1.32 | -7.86 | 1.91 | 0.0004*** | -11.72 |
| Occipital | -2.09 | 1.23 | 0.50 | 3.66 | -4.00 | 1.92 | 0.20 | -7.03 |
| Prefrontal | -2.30 | 1.02 | 0.19 | 1.31 | -3.37 | 2.11 | 0.47 | -4.71 |
| Temporal | -0.88 | 1.10 | 0.98 | 3.79 | -2.87 | 2.17 | 0.66 | -4.40 |
| OFC | -2.15 | 1.56 | 0.66 | 3.61 | 1.77 | 2.60 | 0.98 | 2.85 |
| ACC | -0.56 | 1.48 | 0.99 | 0.77 | -3.47 | 2.54 | 0.63 | -4.55 |
| PCC | -2.89 | 2.90 | 0.92 | 3.60 | -6.04 | 4.36 | 0.62 | -7.40 |
| Insula | -1.18 | 1.39 | 0.96 | 1.76 | -2.62 | 2.38 | 0.82 | -3.66 |
| Putamen | -1.54 | 1.31 | 0.83 | 2.86 | -1.30 | 2.22 | 0.99 | -2.38 |
| Caudate | -1.92 | 1.31 | 0.62 | 4.19 | 0.65 | 2.10 | 1.00 | 1.45 |
| Thalamus | -1.10 | 2.13 | 0.99 | 2.11 | 1.21 | 3.62 | 1.00 | 2.47 |
| Hippocampus | -0.93 | 1.25 | 0.98 | 1.79 | 4.96 | 2.68 | 0.31 | 9.36 |
| Amygdala | -0.56 | 1.11 | 0.99 | -1.12 | -2.46 | 3.06 | 0.96 | -5.07 |
| **ICA diameter** | **-0.001** | **0.05** | **0.99** | **-0.02** | **-0.39** | **0.12** | **0.007\*\*** | **-10.05** |

Expected change in outcome measures (cerebral blood flow and internal carotid artery diameter) as estimated by the Linear Mixed Effects model. Ketanserin effect is presented on the left side of the table, while the drug-x-time interaction effect for psilocybin versus ketanserin is shown on the right side. $\beta$ represents the estimated change between pre-drug and +80 minutes post-drug administration. $\Delta\%$ denotes the estimated percent change for ketanserin (left) and the estimated relative percent change of psilocybin versus ketanserin (right). $P_{FWER}$ indicates family-wise error rate corrected p-values, except for Global CBF and ICA diameter, which are uncorrected. CBF measures are reported for global and regional areas. Brain regions include OFC (Orbitofrontal cortex), ACC (Anterior cingulate cortex), and PCC (Posterior cingulate cortex). SE: standard error. Significance levels: \* $p < 0.05$ , \*\* $p < 0.01$ , \*\*\* $p < 0.001$ .

### Contrasting ketanserin and psilocybin effects on CBF and ICA diameter

Contrasting ketanserin and psilocybin effects on global CBF at 80-minutes post-drug administration relative to baseline, we did not see a statistically significant drug-x-time interaction effect (p = 0.14; Table 2 and Figure 4). However, we did find a statistically significant drug-x-time interaction effect on parietal cortex CBF such that the psilocybin-induced decrease in CBF was significantly larger than the effect of ketanserin (ß=-7.86, 95% CI = significant drug-x-time interaction effects on CBF (Table 2). We also observed a significant drug-x-time Figure 4).

**Figure 4:**
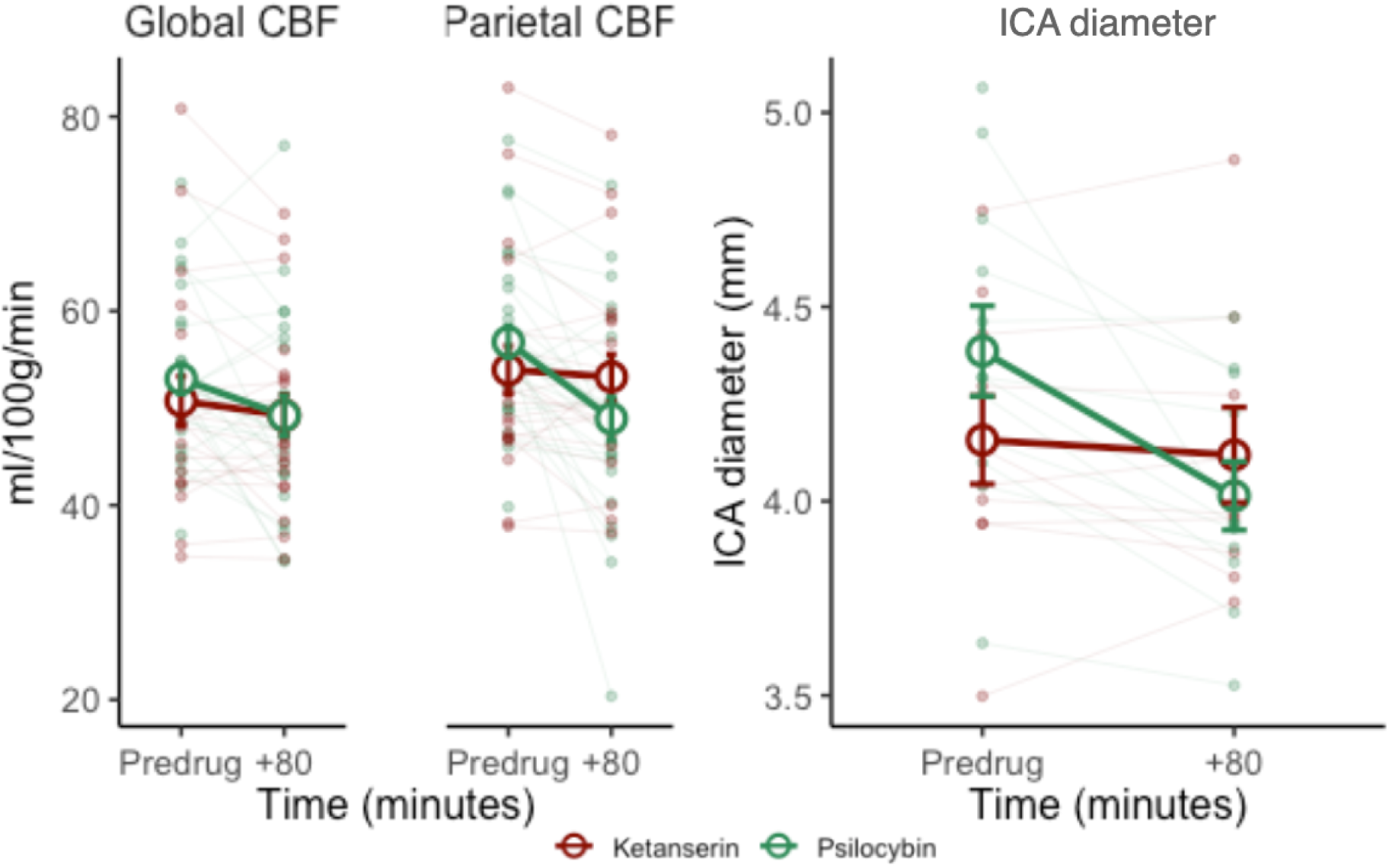
Effects of Ketanserin and Psilocybin on Cerebral Blood Flow and ICA Diameter. Global CBF, Parietal CBF, and ICA diameter shown before and +80-min after administration of psilocybin and ketanserin. Red and green circles and lines represent the ketanserin and psilocybin treatments, respectively. Smaller points denote individual observations, population mean values indicated by larger circles and error bars representing one standard error on the mean.

### Associations between ICA diameter and PPL or SDI

We found that psilocybin intake was associated with a decrease in ICA diameter (Figure 5), with a significant negative association between ICA diameter and both PPL (ß=-0.02, 95% CI = [−0.03, −0.015] mm per μg/L of psilocin, p < 1e-04) and SDI (ß=-0.03, 95% CI = [−0.038, −0.014] mm per unit SDI, p < 1e-04). This corresponds to a 10.5% decrease in ICA diameter at peak PPL and a 6.6% decrease at peak SDI.

**Figure 5.**
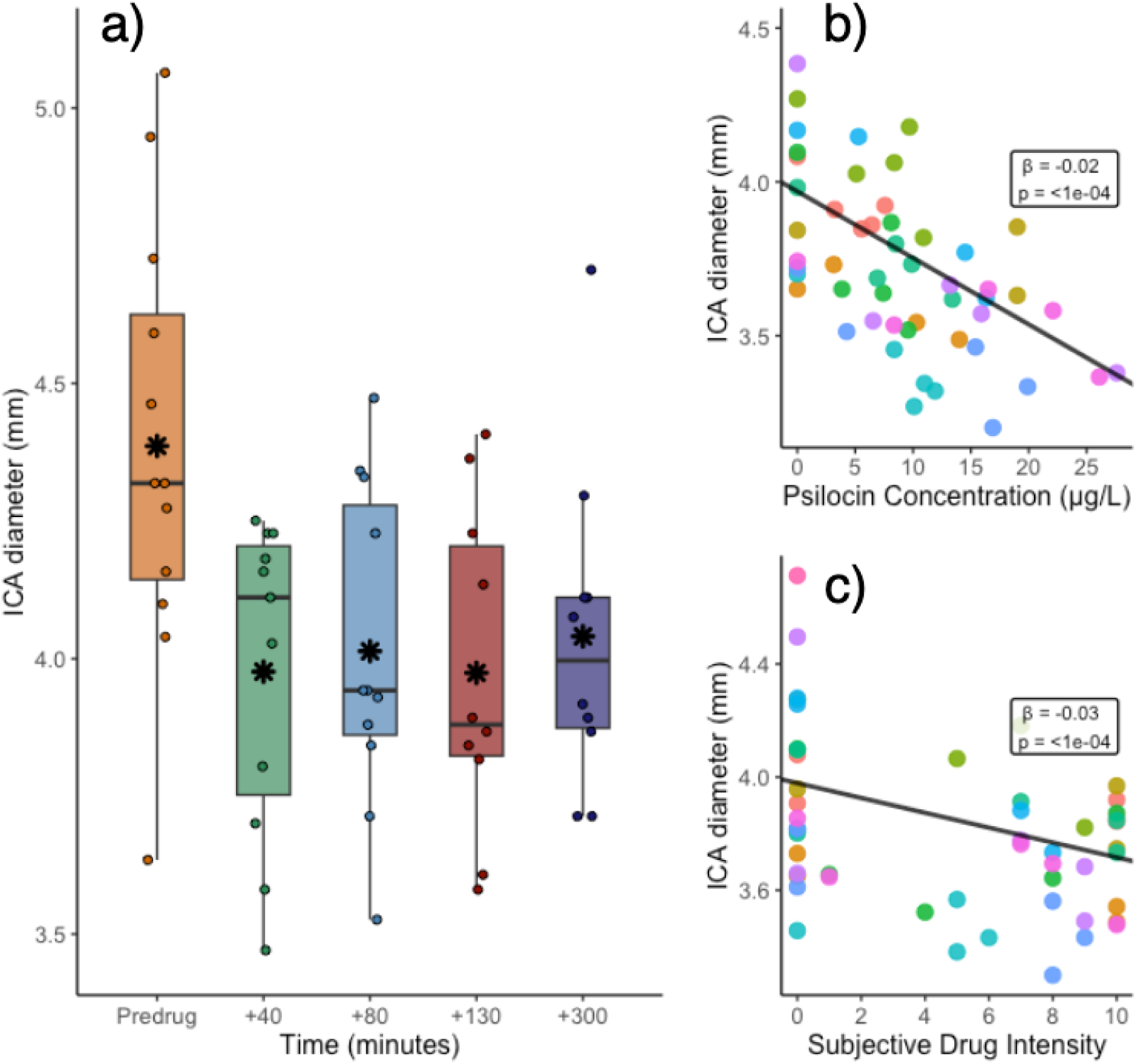
Associations between internal carotid artery (ICA) diameter and plasma psilocin level (PPL) and subjective drug intensity (SDI). **a)** ICA diameter changes over time following psilocybin administration. Box plots show distribution at each time point, with individual measurements as dots. Asterisks indicate means. b) Association between ICA diameter and PPL. Each colour represents a unique individual. c) Association between ICA diameter and SDI. Associations with ICA diameter are adjusted for age, sex.

## Discussion

Here, we evaluated and contrasted the acute effects of psilocybin and ketanserin, drugs with agonist and antagonist effects at the 5-HT2AR, respectively, on global and regional CBF in healthy individuals. Psilocybin intake was associated with up to an 11.6% reduction in global CBF; we show here for the first time that this change is significantly associated with both PPL and SDI, which we have previously shown to be tightly related to brain 5-HT2AR occupancy,^45^ associated with changes in BOLD functional network connectivity^46^, occurrence of dynamic brain states^49^, and certain forms of brain-entropy.^48^ Additionally, we observed a significant decrease in ICA diameter following psilocybin administration, with a 10.5% reduction at peak PPL, highlighting the physiological effect of psilocybin on cerebral vasculature. Neither CBF nor ICA diameter were significantly changed following ketanserin, indicating that 5-HT2AR antagonism does not affect CBF.^60^ In the parietal cortex, psilocybin intake was associated with a statistically significantly larger decrease in CBF compared to that of ketanserin. These findings reinforce an acute modulation of CBF by psilocybin across many brain regions and directly map these changes onto acute pharmacological and subjective effects.

We provide the first evidence that both PPL and SDI, acquired at the time of each individual scan session, are significantly negatively correlated with global and regional CBF, reinforcing a close link between the acute effects of psilocybin, associated subjective effects, and CBF. Our observation that CBF is reduced following psilocybin administration is somewhat consistent with previous studies, but certain discrepancies are noteworthy.^40–42^ In a smaller sample of 15 healthy participants, Carhart-Harris and colleagues reported negative associations between perceptual effects of psilocybin and CBF in the ACC, PCC and thalamus.^40^ We observed similar negative associations between PPL and SDI and CBF within the ACC and PCC, regions often the focus of psychedelic resting-state fMRI literature stemming from being elements of the default-mode network and regions with high 5-HT2AR density.^38,61–63^ However, we did not observe significant associations between PPL nor SDI and thalamus CBF (see Table 1), the eponymous brain region for the thalamic gating model of psychedelic action.^64,65^ Our observation of a limited effect in the thalamus is seemingly consistent with a study by Lewis and colleagues, who report decreased CBF in 58 individuals, i.e., that which they report as “global CBF” or “gCBF”, in large clusters distributed across many brain regions including seemingly only a small subsection of only the left thalamus.^41^ Lewis and colleagues reported lower CBF following psilocybin administration within many regions, including the frontal, parietal, temporal, and occipital cortices, as well as the ACC. Although these observations are consistent with ours, they also reported significant reductions in CBF in the insula, caudate, putamen and amygdala.

We observed a statistically significant negative association between CBF and SDI (but not PPL) in the putamen, with numerically similar but non-significant associations in other subcortical regions. Although Lewis and colleagues reported lower CBF in the hippocampus and thalamus, these effects appear to be confined to relatively smaller subsections, which somewhat aligns with our findings of non-significant associations in these areas. There are notable sources of heterogeneity between the studies, which may explain the discrepant regional results: 1) Carhart-Harris and colleagues administered psilocybin intravenously, whereas Lewis and colleagues and we administered psilocybin perorally, i.e., as it is currently administered in clinical trials, ^3^ 2) Carhart-Harris and colleagues estimated pulsed ASL, whereas Lewis and colleagues performed pcASL, as did we. Approximately half of our data were acquired with a 2D single-PLD sequence, similar to the sequence used by Lewis and colleagues, whereas the other half of our data were acquired with a 3D multi-PLD sequence. Nevertheless, despite regional and magnitude differences, our observation that global and regional CBF is reduced following psilocybin is generally consistent with the previous literature, which we show is significantly correlated with PPL and SDI.

Interestingly, we observed more brain regions showing a significant association between CBF and SDI than PPL and with higher statistical significance (see Table 1). This is qualitatively similar to what we observed when evaluating associations with resting-state functional connectivity in a subset of the current included participants (see Figure 3 from cit.^46^). Our estimated maximal change in CBF, i.e., model estimated decrease at peak observed PPL and SDI, was similar in most neocortical regions i.e. temporal, parietal and occipital, in the range: [−11% to −15.5%], but diverged in deeper brain structures, where the effect of PPL was in the range: [+1% to −13.9%] and SDI in the range [−5.2% to −14.5%]. SDI scores are more bimodally distributed than PPL, which may affect related statistical associations. Notably, 5-HT2AR occupancy by psilocin is related non-linearly to plasma psilocin levels.^45^ If we assume that most of the acute neural effects of psilocybin are due to 5HT2AR agonism, then mapping linear relations between plasma drug levels and neural changes, e.g., CBF, may inadequately capture the true relation between these variables. This may contribute to our observed differences in estimated associations with PPL and SDI; how best to model these associations should be evaluated further in future studies.

Although CBF was numerically decreased across brain regions following ketanserin administration, this effect was small, and not statistically significant (Table 2). Similarly, ketanserin produced negligible perceptual effects. Our lab previously showed with [11C]Cimbi-36 PET that 20 mg ketanserin produces ∼65-75% 5-HT2AR occupancy 1-3 hours after oral administration, near to its estimated maximal occupancy (i.e, 77%)^66^ and the maximal occupancy reported for other 5-HT2AR antagonists/inverse agonists.^49,67,68^ The observation that 5-HT2AR blockade with ketanserin at this occupancy level does not substantively affect CBF suggests that endogenous 5-HT2AR signalling at rest plays a limited role in regulating CBF. Directly comparing effects of psilocybin and ketanserin on CBF, we observed a statistically significant drug-x-time interaction effect within the parietal cortex such that the psilocybin-related decrease was greater (i.e., more negative) than the ketanserin-related change. The parietal cortex is the region wherein we observed the most pronounced negative association between CBF and both PPL and SDI, i.e., a decrease in CBF of ∼15% at peak drug and experience level. The parietal cortex contains regions involved in higher-order association networks including the default mode and frontoparietal networks, alterations in the functional connectivity of which have been linked to acute psychedelic effects.^40^ Although other regions in those networks showed directionally similar drug-x-time interaction effects, they were not statistically significant. We interpret the associations with PPL and SDI as supportive of a substantial psilocybin effect on CBF.

It is not immediately straight-forward how to reconcile the generally observed decrease in CBF with previous psilocybin studies evaluating effects on cerebral metabolic rate of glucose (CMRglc) with [18F]-FDG PET. One previous study reported numerically decreased global but both increased (e.g., anterior cingulate) and decreased (e.g., thalamus) regional CMRglc following psilocybin administration in the context of a word repetition/association task.^69^ However, a second study that did not include any such task reported a global increase in CMRglc of ∼10-25%, most markedly in the frontal, anterior cingulate gyrus, and temporomedial cortex.^44^ This discrepancy is somewhat unexpected considering that CMRglc and CBF are normally tightly coupled.^70,71^ However, decoupling between CBF and CMRglc has been reported previously, e.g., caffeine has been shown to decrease CBF ^72–75^ and increase energy metabolism,^76,77^ and previous studies have speculated that psilocybin-induced reductions in CBF are not due to effects of stimulation of the neuronal 5-HT2ARs, but other receptor targets and direct effects on vascular tone.^40,41^ One possible explanation for the discrepancy between the current results and the previous reports of increased CMRglc following psilocybin ingestion could be the agonist activity of psilocybin at the inhibitory serotonin 1A receptor (5-HT1AR). The 5-HT1AR is expressed in various brain regions and acts both as an auto- and hetero-receptor and has, upon stimulation, been shown to lead to reduced cerebral blood volume in animal models.^18,78,79^

We also observed that ICA diameter was significantly negatively associated with both PPL and SDI, the first human *in vivo* evidence that psilocybin induces cerebral vasoconstriction. Our psilocybin observations are consistent with previous studies showing 5-HT2AR is expressed in the peripheral vasculature,^29^ where it induces vasoconstriction,^30,32,80,81^ and preclinical research shows 5-HT-induced vasoconstriction in the carotid arteries of rats.^82,83^ This highlights an important consideration for studies estimating CBF effects of 5-HT2AR agonist drugs: pcASL is inversely related to carotid blood flow velocity in a non-linear fashion, with the effects being most pronounced for increases in flow velocity.^84,85^ A psilocin-induced vasoconstriction could reduce the amount of arterial blood being labelled, thereby underestimating CBF. Additionally, both preclinical and clinical studies have shown that psilocybin can induce mild hyperthermia,^86,87^ which in animals induces carotid artery constriction.^88^ Based on our current observation and previously suggested best practice,^89^ future studies of psychedelic effects on CBF should measure vasoactive effects with, e.g., TOF angiography and phase-contrast mapping to directly measure ICA diameter and blood flow velocity, respectively. This will assist the interpretation of CBF effects and provide critical insights into the hemodynamic changes induced by these drugs.

Multimodal imaging techniques, such as simultaneous [18F]-FDG PET and pcASL, may also help resolve whether the apparent discrepancy between CMRglc and CBF reflects a decoupling of CBF from energy metabolism following psilocybin administration. Future studies using more selective 5-HT2AR agonists (such as the phenethylamine psychedelics^90^) and 5-HT1AR antagonists could help disentangle various sources of CBF alterations in CBF studies using polypharmacological drugs like psilocybin and ketanserin. These approaches would provide more detailed information about the relation between psilocybin, 5-HT2AR, and hemodynamic responses, clarifying the contributions of neuronal and non-neuronal factors to associated signals that are a common source of interest.^3^

One of the previous studies that evaluated acute psilocybin effects on CBF considered two analysis strategies, one of which involved adjusting for global CBF.^41^ This was motivated in part by a previous study advocating for this strategy.^91^ We did not apply this adjustment because our repeated measures model relates within individual changes in CBF to PPL or SDI, which we view as particularly desirable for assessing associated drug effects. It is also important to note that although we observed some grey-matter weighted CBF estimates outside the typically expected range of 40-80 ml/100g/min,^89^ most of our scans were within this range.

Our study is not without its limitations. Employing a double-blind design could have limited potential biases compared to our single-blind design. Notwithstanding, maintaining participant blinding in psychedelic studies is very difficult due to the intense subjective effects.^4,11,17,95–97^ Brain imaging data were collected across two scanners with distinct pcASL sequences (2D vs. 3D acquisition protocol) because MR1 became unavailable during the study and the MR1 sequence was not available on the MR2 scanner. These represent hardware and software sources of variation, which we adjust for by including scanner as a variable in our statistical models. This may potentially limit our ability to resolve drug-related effects. As we have discussed previously,^12^ hysteresis effects, i.e., different associations during the ascent and descent phases, cannot be ruled out; we do not have sufficient data to directly estimate hysteresis effects. We corrected for motion and excluded participants with excessive motion, yet it may remain a confound as head motion is increased while on psychedelics.^40,95^ Psilocybin has been found to increase body temperature, blood pressure, and heart rate. ^87^ Although changes in respiration, cardiac cycle, and end-tidal carbon dioxide during psilocybin sessions could potentially influence CBF, one previous study found that these factors did not significantly alter the CBF analysis. ^40^

Overall, we find that CBF decreases acutely following psilocybin administration and it is negatively associated with PPL and SDI. By contrast, ketanserin did not significantly affect CBF. ICA diameter significantly decreased following psilocybin, highlighting the need to resolve physiological mechanisms important for interpreting psychedelic effects on hemodynamic related signals. These findings add to our understanding of the neurobiological effects of psychedelic drugs and may have implications for their potential therapeutic uses in the treatment of brain disorders.

## Data Availability

The MATLAB and R code used to generate the results presented in this study can be made available upon request. The datasets generated and/or analysed during the current study can be made available upon completion of a formal data-sharing agreement.

## Acknowledgments

We gratefully acknowledge the work of MRI assistants, Agnete Dyssegaard and Arafat Nasser for biobank management, and Oliver Overgaard-Hansen and Vibeke Dam for assistance in guiding psilocybin interventions. We thank Lone Freyr, Gerda Thomsen, Svitlana Olsen, Peter Jensen and Dorthe Givard for technical/administrative assistance. We also acknowledge the BAFA laboratory, University of Chemistry and Technology and the National Institute of Mental Health (Prague, Czech Republic) for production of psilocybin and Glostrup Apotek (Glostrup, Denmark) for encapsulation. We would like to thank Professor Fernando Calamante from the University of Sydney for his manuscript feedback related to pcASL imaging. We would like to thank the Department of Radiology at Rigshospitalet for MR1 scanner access. We would like to thank the Kirsten & Freddy Johansen (KFJ) Foundation for funding the MR2 scanner.

## Author contributions

KL contributed to data analysis, visualisation, interpretation and wrote the first draft of the manuscript. UL contributed to data analysis. BO and DE-WM both contributed to data analysis and interpretation. SA contributed to guiding psilocybin interventions, and participant preparation and integration. AJ contributed to the protocol and data acquisition. DSS conceptualised the main psychological outcome of the study (SDI), study settings, and guided psilocybin interventions, participant preparation and integration. PMF, GMK and MKM, designed the study, wrote the protocol, and contributed to data collection, analysis and interpretation. All authors contributed substantially to interpretation and discussion of the study results, to critical review of the submitted manuscript, approval of the final version, and agreed to be both personally accountable for their own contributions and willing to participate in resolving any questions that may arise regarding the present paper.

## Supplementary Materials

**Supplementary Figure S1.**
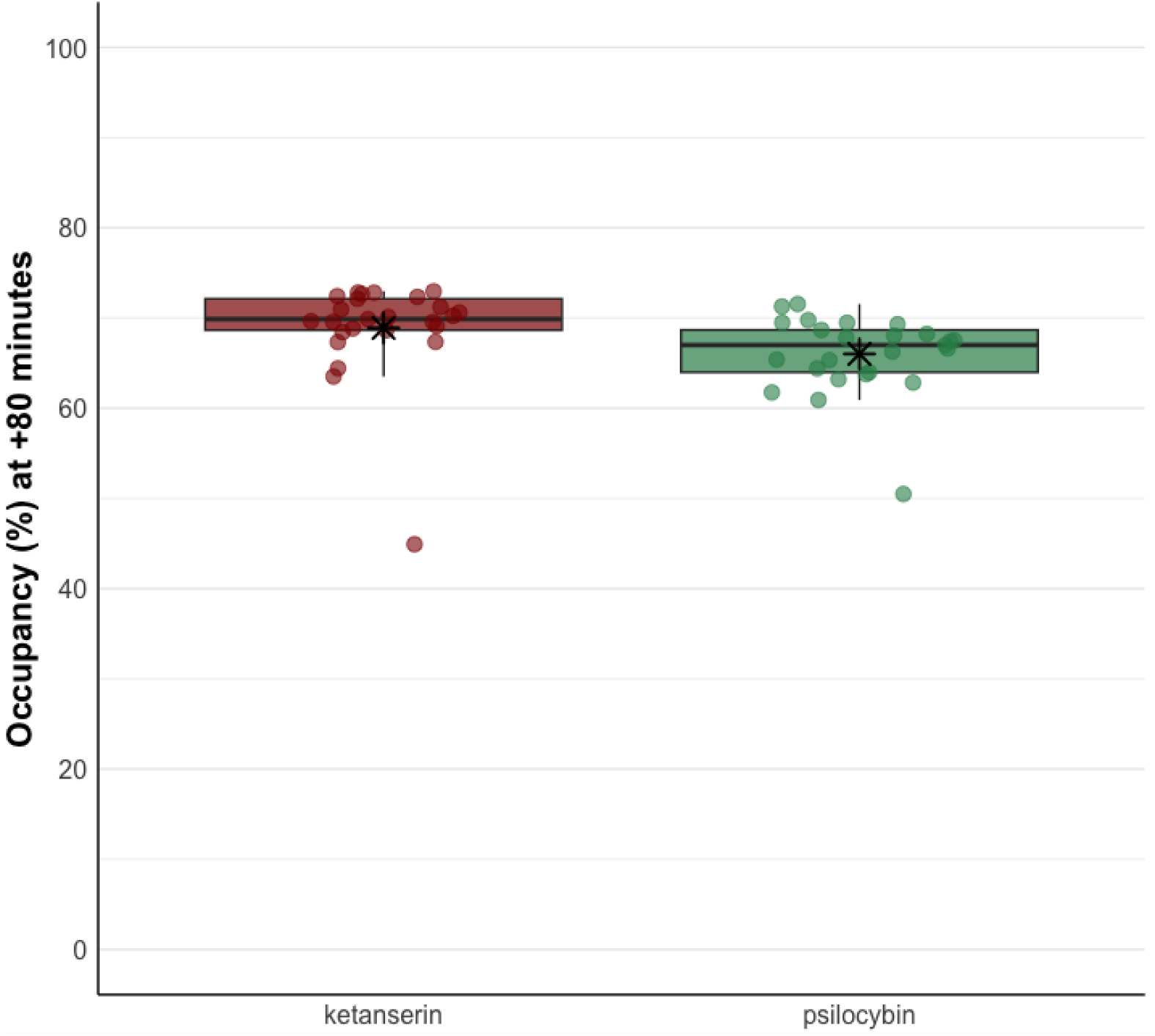
Estimated receptor occupancy for ketanserin and psilocybin at +80 minutes post-drug. This plot shows the estimated receptor occupancy (%) at 80 minutes after drug administration for ketanserin (red) and psilocybin (green). Each dot represents an individual participant’s estimated occupancy. The box plots display the median (horizontal line), interquartile range (box), and 1.5 times the interquartile range (whiskers). The black asterisks indicate the mean occupancy for each drug. Occupancy was estimated using plasma concentrations and the Emax model derived from our previous work.

**Supplementary Figure S2.**
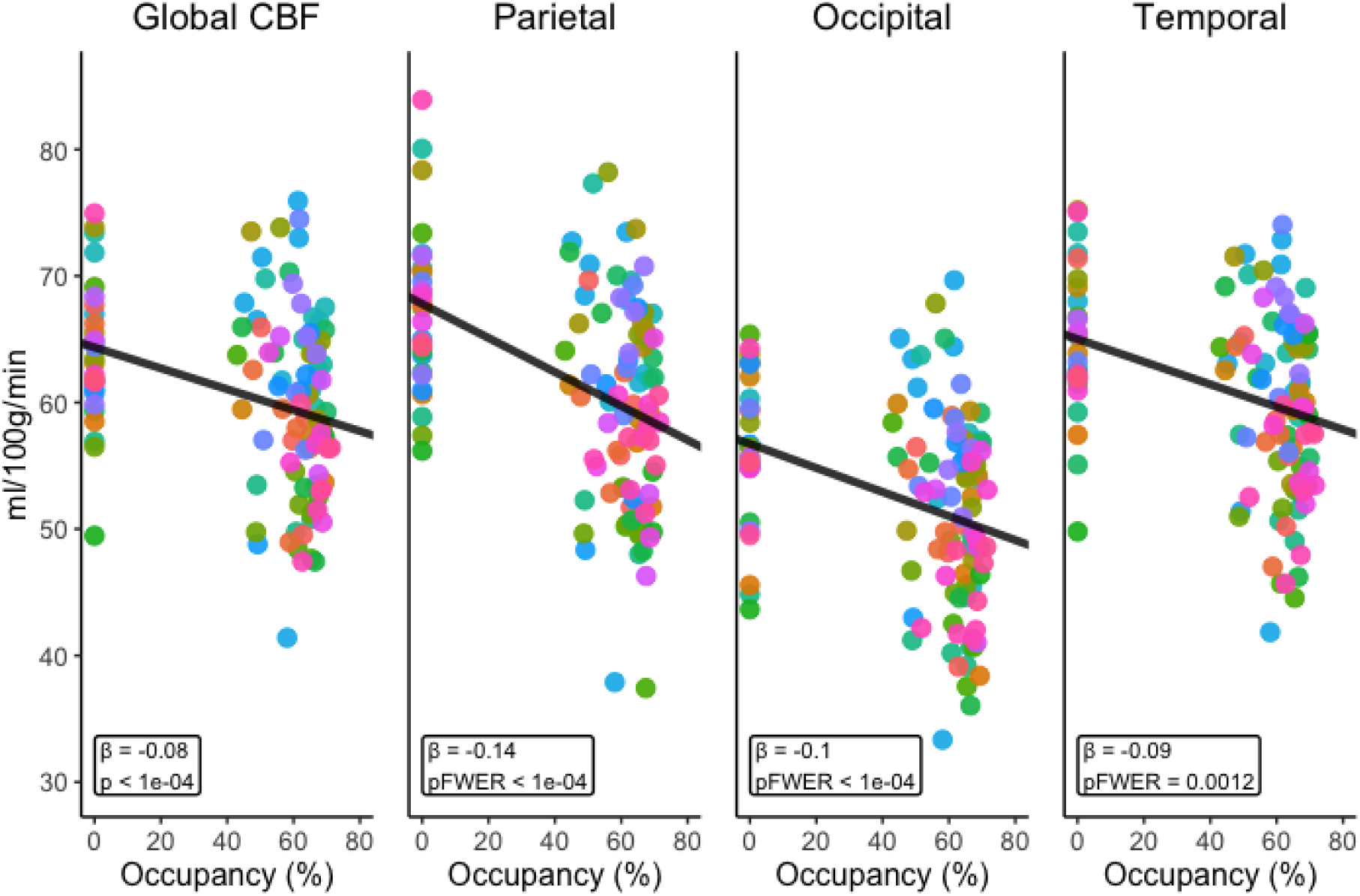
Associations between regional cerebral blood flow (CBF) and estimated receptor occupancy. This figure shows the relationships between estimated receptor occupancy (%) and CBF (ml/100g/min) in four brain regions: Global CBF, Parietal, Occipital, and Temporal cortices. Each panel represents a different brain region, with estimated receptor occupancy on the x-axis and CBF on the y-axis. The black line in each panel indicates the fitted linear relation between occupancy and CBF, adjusted for age, sex, and scanner. Each coloured dot represents a single measurement from a unique participant. The β values shown in each panel represent the slope of the fitted line, indicating the change in CBF per unit increase in occupancy. The p or pFWER values indicate the statistical significance of the association, with pFWER values used for regional analyses to account for multiple comparisons.

---

**Table S1.** Association between CBF and 5-HT2ARoccupancy.

| MEASURE | 5-HT2AR OCCUPANCY |  |  |  |
| --- | --- | --- | --- | --- |
| | $\beta$ | SE | P<sub>FWER</sub> | $\Delta\%$ |
| **Global CBF** | **-0.38** | **0.09** | **<1e-04\*\*\*** | **-9.88** |
| Parietal | -0.14 | 0.02 | <1e-04*** | -15.20 |
| Occipital | -0.10 | 0.02 | <1e-04*** | -12.84 |
| Prefrontal | -0.07 | 0.03 | 0.022* | -8.06 |
| Temporal | -0.09 | 0.02 | 0.001** | -10.52 |
| OFC | -0.02 | 0.04 | 0.99 | -2.41 |
| ACC | -0.07 | 0.03 | 0.049* | -7.23 |
| PCC | -0.12 | 0.04 | 0.017* | -10.62 |
| Insula | -0.06 | 0.03 | 0.20 | -6.85 |
| Putamen | -0.04 | 0.02 | 0.29 | -5.79 |
| Caudate | -0.02 | 0.02 | 0.85 | -3.71 |
| Thalamus | 0.02 | 0.04 | 0.99 | 3.44 |
| Hippocampus | 0.04 | 0.03 | 0.42 | 5.90 |
| Amygdala | -0.06 | 0.03 | 0.24 | -8.97 |

Expected change in outcome measure (cerebral blood flow) for a unit change in 5-HT2AR occupancy as estimated by the linear mixed model. $\beta$ represents the estimated change. $\Delta\%$ denotes the estimated percent change for occupancy = 76.6% (Emax) versus occupancy = 0%. P<sub>FWER</sub> indicates family-wise error rate corrected p-values, except for Global CBF, which is uncorrected. CBF measures are reported for global and regional areas. Brain regions include OFC (Orbitofrontal cortex), ACC (Anterior cingulate cortex), and PCC (Posterior cingulate cortex). SE: standard error. Significance levels: \* p < 0.05, \*\* p < 0.01, \*\*\* p < 0.001.

